# Patient characteristics associated with participation in cardiorespiratory exercise during stroke rehabilitation: a multisite observational cohort study

**DOI:** 10.64898/2026.04.01.26349980

**Authors:** Augustine J. Devasahayam, Ada Tang, Yuan Zhong, Osvaldo Espin-Garcia, Sarah Munce, Kathryn M. Sibley, Elizabeth L. Inness, Avril Mansfield

## Abstract

**Objectives:** Among individuals attending stroke rehabilitation, we aimed to determine the proportion who participated in cardiorespiratory exercise, identify patient characteristics predicting participation, and describe exercise characteristics.

**Design, setting, and participants:** This was an observational cohort study involving all patients admitted to four stroke rehabilitation centres in Ontario, Canada, during March or October 2019, or over 12 months starting in 2021.

**Main measures:** Patient characteristics extracted during chart review included age, sex, marital status, employment status, date of stroke, time post-stroke at admission, length of stay for rehabilitation, past medical history that could affect exercise participation, Functional Independence Measure, Functional Ambulation Category, mobility aid use, Chedoke-McMaster Stroke Assessment, Montreal Cognitive Assessment, National Institutes of Health Stroke Scale, and details describing cardiorespiratory exercise completed.

**Results:** 40.1% of stroke patients participated in cardiorespiratory exercise, with 26.4% having it included in their treatment plan. Diagnosed cardiac disease (OR=0.74), poor left ventricular function (OR=0.09), history of mental health conditions (OR=0.69), lower functional ambulation ability (OR=0.74), and wheelchair use at rehabilitation admission (OR=0.46) were associated with lower odds of participating in cardiorespiratory exercise after stroke (p-values<0.05). Use of a walker or rollator at rehabilitation admission (OR=3.22), having a cardiorespiratory exercise goal (OR=2.13), and longer lengths of stay (OR=1.01) were associated with higher odds of participating in cardiorespiratory exercise after stroke (p-values<0.05). Only 1.5% of patients (N=9/601) who participated in cardiorespiratory exercise completed it with recommended intensity and duration.

**Conclusion:** Improving participation in cardiorespiratory exercise during stroke rehabilitation may require addressing cardiovascular, mental health, and mobility-related barriers.

## BACKGROUND

People who have experienced a stroke often face challenges related to low cardiorespiratory fitness, which can significantly impact their ability to perform activities of daily living.^1–3^ Cardiorespiratory fitness refers to the capacity of the cardiorespiratory system to transport oxygen to the muscles during prolonged physical activity.^4,5^ Low cardiorespiratory fitness can lead to fatigue, decreased endurance, and difficulty in performing tasks that require physical exertion, such as walking, climbing stairs, dressing and bathing.^6^ Stroke survivors may experience low cardiorespiratory fitness due to various factors, such as decreased mobility, muscle weakness, and impaired cardiovascular function.^6^

Prescribing cardiorespiratory exercise during the subacute stage of stroke in inpatient rehabilitation can increase cardiorespiratory fitness post-stroke.^7,8^ About 78% of Canadian stroke rehabilitation programs^9^ and 76% of physiotherapists who treat people with stroke^10^ report prescribing cardiorespiratory exercise. However, across 5 sites in Ontario, Canada, only 6-42% of people admitted to stroke rehabilitation participated in a cardiorespiratory exercise^7,8^ when an individualized exercise program was delivered in a group setting outside of routine care.^8,11^ Therefore, it is possible that the number of patients participating may be even lower in settings where there are additional barriers to implementation, including limited staffing, inadequate space or equipment, and healthcare professionals’ limited knowledge and confidence in delivering stroke-specific cardiorespiratory exercise programs.^12^ Institutional priorities and variability in clinician training may also contribute to inconsistent program uptake across facilities.^9,13,14^ Therefore, it is necessary to determine the proportion of stroke patients who engaged in cardiorespiratory exercise during routine rehabilitation care in both outpatient and inpatient settings across multiple sites.

Canadian physiotherapists who prescribed exercises for patients with stroke reported that individual patient factors, such as severity of neurological condition, patients’ rehabilitation goals, comorbidities, mobility level, current fitness level, fall risk, pre-stroke physical activity history, results of an exercise test, age, and sex, were considered in determining whether cardiorespiratory exercise should be prescribed.^10^ Additionally, physiotherapists at one Canadian centre with a structured cardiorespiratory exercise program for stroke patients identified cardiovascular risk, fatigue, and cognitive impairment as major challenges in prescribing and implementing cardiorespiratory exercise.^15^ These factors represent physiotherapists’ perceptions, but their actual impact on stroke patients’ participation in cardiorespiratory exercise remains unclear. Previous research found that patient characteristics such as age, presence of cardiac disease, potential cardioembolic sources, and arthritis, differed between those enrolled and not enrolled in a structured cardiorespiratory exercise early after stroke.^7^ However, these findings were based on a retrospective review with a limited sample in a single site. Determining the patient characteristics that are associated with participation in cardiorespiratory exercise after stroke is critical for the success of stroke rehabilitation programs. Analysis of patient characteristics predicting participation in cardiorespiratory exercise across multiple rehabilitation centres will provide a comprehensive understanding of the factors that influence participation in cardiorespiratory exercise during stroke rehabilitation. Furthermore, it remains unknown if these cardiorespiratory exercise prescriptions adhered to evidence-based guidelines,^16,17^ making it difficult to assess the effectiveness of these programs.

## OBJECTIVES

The primary aim of this study was to determine the proportion of people with stroke who participated in cardiorespiratory exercise during rehabilitation in four Canadian sites. As data collection for this study happened during the coronavirus 2019 (COVID-19) pandemic, we explored trends in cardiorespiratory exercise participation before and after the onset of the pandemic. The secondary aims were to determine patient characteristics that predicted participation in cardiorespiratory exercise after stroke, both for all patients and for inpatients separately, and to describe characteristics of cardiorespiratory exercise completed during stroke rehabilitation in relation to evidence-based clinical practice guidelines, including average session attendance, use of standardized exercise testing pre- and/or post-participation, type of equipment used, and whether the prescribed exercise met recommended dosage (i.e., moderate intensity for ≥20 minutes per session).^16,17^

## METHODS

### Study design

This was a multisite observational cohort study. The study was approved by the research ethics boards of the University Health Network (protocol number: 20-5695), Sunnybrook Health Sciences Centre (protocol number: 3605), and Hamilton Integrated Research Ethics Board (protocol number: 11523). This study is reported using the STROBE guidelines for observational cohort studies.^18^

### Setting and participants

The four participating sites were urban academic centres with dedicated stroke rehabilitation services in Ontario, Canada. Sites provided both inpatient and outpatient stroke rehabilitation services, except one, which offered outpatient services only. Patients were included in the chart review if they were admitted to the stroke rehabilitation program in either March or October 2019 (pre-COVID-19 pandemic), or during a 12-month period starting in 2021 (post-COVID-19 pandemic; see Results for specific dates), to ensure comprehensiveness in evaluating the patient characteristics related to rehabilitation services before and after the onset of COVID-19. All patients admitted to the stroke rehabilitation program were eligible, and no *a priori* exclusion criteria were applied. A waiver of patient consent for chart review was approved by each site’s research ethics board. However, patients in the post-pandemic cohorts were informed of the study and had the opportunity to opt out of chart review.

### Descriptors

Patient characteristics extracted during chart review were: age, sex, marital status, employment status, date of stroke, time post-stroke at admission, length of stay in rehabilitation, past medical history and current co-morbid conditions that could affect exercise participation, Functional Independence Measure (FIM; available for in-patients only),^19^ Functional Ambulation Category (FAC),^20^ mobility aid use, Chedoke-McMaster Stroke Assessment (CMSA),^21^ Montreal Cognitive Assessment (MOCA),^22^ National Institutes of Health Stroke Scale (NIH-SS),^23^ rehabilitation goals, whether the patient had participated in cardiorespiratory exercise during rehabilitation, whether cardiorespiratory exercise was included in their treatment plan, and details of exercise tests and participation (i.e., frequency, intensity, type, duration, and total number of exercise sessions completed).

The Functional Independence Measure (FIM),^19^ which consists of 18 items covering motor and cognitive functions, was used to evaluate physical and cognitive disability in inpatients by measuring their level of independence in daily activities. Each item is scored on a 7-point scale, ranging from 1 (total assistance) to 7 (complete independence), with a total possible score of 126 (91 for motor and 35 for cognitive function). Higher scores indicate greater independence, while lower scores indicate a greater need for assistance. The Functional Ambulation Category (FAC),^20^ a 6-point scale ranging from 0 (unable to walk) to 5 (independent walking on all surfaces), was used to assess walking ability and the level of physical support required for ambulation. Higher scores indicate greater walking independence, while lower scores indicate a higher need for assistance during ambulation.

The Chedoke-McMaster Stroke Assessment (CMSA),^21^ a seven-stage measure of motor recovery, was used to assess motor function in the arm, hand, leg, and foot following stroke. The stages range from flaccid paralysis (Stage 1) to normal movement (Stage 7), with higher scores indicating greater motor recovery and lower scores indicating more severe impairment. The Montreal Cognitive Assessment (MOCA),^22^ a screening tool for cognitive impairment, was used to assess visuospatial/executive function, naming, memory, attention, language, abstraction, delayed recall, and orientation. Scores range from 0 to 30, with higher scores indicating better cognitive function and lower scores indicating greater impairment. The National Institutes of Health Stroke Scale (NIH-SS),^23^ a tool for evaluating the severity of stroke symptoms, was used to assess neurological function across 11 domains: level of consciousness, best gaze, visual fields, facial palsy, motor arm, motor leg, limb ataxia, sensory, best language, dysarthria, and extinction and inattention. Scores range from 0 to 42, with higher scores indicating more severe impairment and lower scores indicating less neurological deficit.

### Data sources

Data were obtained from medical charts. Demographic variables (age, sex, marital status, and employment status) and FIM scores were collected from administrative records at the time of admission to stroke rehabilitation. Time post-stroke at admission was obtained from administrative records by calculating the difference between the stroke onset date and the admission date. The length of stay in rehabilitation was determined based on the total number of days from admission to discharge. Past medical history and co-morbid conditions that could impact exercise participation, including cardiovascular disease, musculoskeletal disorders, cancer, metabolic syndromes, mental health conditions, respiratory diseases, and neurological conditions, were extracted from physician notes. The FAC was scored by researchers based on physiotherapists’ and/or occupational therapists’ notes regarding gait assessment/functional mobility. CMSA scores were obtained from physiotherapists’ assessments. MOCA scores were obtained from occupational therapist or physician assessment. Mobility aids used and NIH-SS scores were obtained from various sources, including physician notes, therapist documentation, and other standardized reports at the time of admission to stroke rehabilitation.

All rehabilitation goals related to physical function were extracted verbatim; the research team determined that goals were related to cardiorespiratory exercise if the goal was to improve cardiorespiratory fitness (e.g., “to improve endurance”), or to return to participation in sports or exercise (e.g., hiking, cycling). Participation in cardiorespiratory exercise and details of exercise participation were determined from physiotherapists’ assessments, treatment plans, and treatment documentation (e.g., progress notes, exercise logs). If the physiotherapist treatment plan included cardiorespiratory exercise (exercise intervention plans contained terms such as, endurance, aerobic fitness, for example) and cardiorespiratory exercise was documented in exercise logs/progress notes, then patients were classified as “prescribed”. If cardiorespiratory exercise was not included in the treatment plan, but exercise logs or progress notes suggested that patients completed cardiorespiratory exercise, then they were classified as “incidental”. Cardiorespiratory exercise test results (e.g., maximal cardiopulmonary exercise test (CPET), graded sub-maximal test, 6-minute walk test, or 2-minute walk test) were extracted from related assessment forms. Frequency of exercise (number of sessions per week), intensity (e.g., rate of perceived exertion, target heart rate), type of activity (e.g., treadmill walking, cycling), and duration (minutes per session) were extracted from physiotherapists’ notes. The total number of cardiorespiratory exercise sessions was obtained from exercise logs/progress notes. Exercise intensity was categorized as moderate^16^ if any of the following criteria were met: an RPE of ≥3 on a 10-point scale,^24^ an RPE of ≥11 on a 20-point scale,^25^ a heart rate of ≥55% of maximum or age-predicted maximum, or a qualitative description indicating “moderate intensity” exercise. Exercise duration was expressed as a range with an upper and lower limit. We determined if exercise participation met evidence-based clinical practice guidelines,^16,17^ i.e., moderate intensity exercise for a minimum of 20 minutes per session.

### Study size

Based on admission rates for stroke rehabilitation at the participating sites in the previous years, the expected number of available charts across all sites combined was 205 for the pre-COVID cohort (TRI-UC: 75, TRI-RC: 23, SJR: 57, HHS: 50), and 1,230 for the post-COVID cohort (TRI-UC: 450, TRI-RC: 140, SJR: 340, HHS: 300). We expected 6-42% of patients to participate in cardiorespiratory exercise during stroke rehabilitation based on previous work.^7,8,11^ The performance of the multivariable models in predicting participation in cardiorespiratory exercise during stroke rehabilitation was assessed via the area under receiver operating characteristic (AUROC) curves. A sample size of at least 615 participants was required to achieve >80% statistical power to detect an AUROC of 0.7 (against an AUROC of 0.6 under the null hypothesis), assuming that at least 13% of patients would participate in cardiorespiratory exercise during stroke rehabilitation, at an alpha level of 5%.

### Data analysis

Data analysis was conducted using R (version 4.3.0, R Project for Statistical Computing, Vienna, Austria). The proportion of people with stroke who participated in cardiorespiratory exercise during rehabilitation was calculated considering patients admitted to both in- and out-patient stroke rehabilitation services together. The differences and similarities in clinical profiles between those who did and did not participate in cardiorespiratory exercise during stroke rehabilitation were characterized using parametric (e.g., t-tests, ANOVA) or nonparametric (e.g., Mann-Whitney U test, Kruskal-Wallis test) tests for continuous variables, depending on the data distribution. For categorical variables, Chi-square tests were used to assess the differences. Monte Carlo simulations were performed to assess the reliability of variables under different data distributions and varying levels of missing data, and check the accuracy of the results. Frequency counts and descriptive statistics were used to report exercise prescriptions.

Missing data were imputed using the missForest package, a random forest-based imputation method, under the assumption of missing at random. To determine whether patient characteristics affected participation in cardiorespiratory exercise during stroke rehabilitation, stepwise logistic regression was conducted, including both inpatients and outpatients from all four participating sites. The response variable was participation in cardiorespiratory exercise (yes/no) during stroke rehabilitation. The explanatory variables were patient characteristics collected from chart review: demographic (age, sex), socio-economic (marital status, employment status), admission-related factors (time post-stroke at admission, length of stay in rehabilitation), current co-morbid medical conditions (heart or cardiovascular condition, high blood pressure, musculoskeletal conditions or injuries (including arthritis, osteoporosis, or back problems), cancer of any kind, metabolic conditions (including type 1 or 2 diabetes), mental health conditions, respiratory disease, other neurologic conditions (including concussion and epilepsy), transient ischemic attack, previous stroke, kidney problems), and indicators of physical and cognitive function (FIM, FAC, use of mobility aids, CMSA – leg, foot, arm, hand, MOCA, and NIH-SS score).

The prediction performance of the full sample model was evaluated using 500 iterations of repeated sample splitting. In each iteration, the dataset was randomly split into a 90% training set (n = 1347) and a 10% testing set (n = 150), with model performance assessed using classification metrics, including sensitivity, specificity, and AUROC curve.

A separate analysis was conducted for the inpatient subgroup to identify potential differences in factors influencing participation in cardiorespiratory exercise. The prediction accuracy of the inpatient subgroup model was also evaluated using 500 iterations of repeated sample splitting, with the dataset randomly split into a 90% training set (n = 695) and a 10% testing set (n = 77).

## RESULTS

### Data availability

No patients among the first 154 charts reviewed at one of the study sites completed cardiorespiratory exercise; therefore, partway through data collection we decided to only include a six-month review period for the post-pandemic cohort for this site. There were a total of 1,557 admissions to the stroke rehabilitation programs during the review periods (Table 1). Of these, 60 were excluded from the review: 40 patients opted out, charts could not be located for 19 admissions because they had been requested by another unit or hospital and had not yet been returned to health records department, and one additional inpatient admission was excluded for a patient who was readmitted after experiencing another stroke. Therefore, a total of 1,497 charts were included in the analysis (Tables 1 and 2). A summary of missing values is presented in the supplementary material A.

**Table 1.** Number of charts included in the analysis. Values presented are counts. Patients admitted to the stroke rehabilitation programs between the chart review start and end dates (inclusive) were included in the review.

|  | HHS |  | SJR |  | TRI-RC | TRI-UC |  | Total |
| --- | --- | --- | --- | --- | --- | --- | --- | --- |
|  | In-patient | Out-patient | In-patient | Out-patient | Out-patient | In-patient | Out-patient |  |
| **Pre-COVID** |  |  |  |  |  |  |  |  |
| **Charts reviewed (n)** | 65 | 0 | 56 | 45 | 21 | 43 | 41 | 271 |
| **Post-COVID** |  |  |  |  |  |  |  |  |
| **Admissions (n)** | 151 | 17 | 231 | 226 | 141 | 251 | 269 | 1286 |
| **Excluded (n)** | 0 | 0 | 1 | 14 | 15 | 23 | 7 | 60 |
| **Charts reviewed (n)** | 151 | 17 | 230 | 212 | 126 | 228 | 262 | 1226 |
| **Chart review start date** | 3-Jan-2023 | 1-Feb-2023 | 1-Jun-2022 | 1-Jun-2022 | 1-Feb-2022 | 11-Jan-2021 | 28-Nov 2022 |  |
| **Chart review end date** | 11-Jul-2023 | 1-Sept-2023 | 31-May-2023 | 31-May-2023 | 1-Feb-2023 | 10-Jan-2022 | 27-Nov 2023 |  |
| **Total Included in analysis (n)** | 216 | 17 | 286 | 257 | 147 | 271 | 303 | 1497 |

HHS: Hamilton Health Sciences; SJR: St. John's Rehabilitation; TRI-RC: Toronto Rehabilitation Institute – Rumsey Centre; TRI-UC: Toronto Rehabilitation Institute – University Centre; COVID: Coronavirus Disease; n: Number;

**Table 2.** Prevalence of cardiorespiratory exercise participation among individuals with stroke. Values presented are counts with percentages in parentheses.

|  | HHS |  | SJR |  | TRI-RC | TRI-UC |  | Total |
| --- | --- | --- | --- | --- | --- | --- | --- | --- |
|  | In-patient | Out-patient | In-patient | Out-patient | Out-patient | In-patient | Out-patient |  |
| **Total number participated in cardiorespiratory exercise** |  |  |  |  |  |  |  |  |
| **Pre-COVID** | 2 (3.1) | - | 43 (76.8) | 10 (22.2) | 7 (33.3) | 17 (39.5) | 15 (36.6) | 94 (34.7) |
| **Post-COVID** | 15 (9.9) | 3 (17.6) | 195 (84.8) | 173 (81.6) | 32 (25.4) | 27 (11.8) | 62 (23.7) | 507 (41.4) |
| **Cardiorespiratory exercise included in treatment plan ('Prescribed')** |  |  |  |  |  |  |  |  |
| **Pre-COVID** | 2 (3.1) | - | 15 (26.8) | 5 (11.1) | 5 (23.8) | 13 (30.2) | 15 (36.6) | 55 (20.3) |
| **Post-COVID** | 15 (9.9) | 3 (17.6) | 117 (50.9) | 128 (60.4) | 19 (15.1) | 19 (8.3) | 39 (14.9) | 340 (27.8) |
| **Cardiorespiratory exercise not included in treatment plan ('Incidental')** |  |  |  |  |  |  |  |  |
| **Pre-COVID** | 0 (0) | - | 28 (50.0) | 5 (11.1) | 2 (9.5) | 4 (9.3) | 0 (0) | 39 (14.4) |
| **Post-COVID** | 0 (0) | 0 (0) | 78 (33.9) | 45 (21.2) | 13 (61.9) | 8 (3.5) | 23 (8.8) | 167 (13.6) |

HHS: Hamilton Health Sciences; SJR: St. John's Rehabilitation; TRI-RC: Toronto Rehabilitation Institute – Rumsey Centre; TRI-UC: Toronto Rehabilitation Institute – University Centre; COVID: Coronavirus Disease;

### Participants

There were no statistically significant differences in age, sex, or marital status between individuals who participated in cardiorespiratory exercise after stroke and those who did not (p-values, 0.47, 0.58, 0.54, respectively; Table 3). The employment status of patients was significantly associated with participation in cardiorespiratory exercise after stroke (p = 0.025; Table 3). Specifically, a higher proportion of retired patients participated in exercise (58%) compared with those who did not participate (51%), whereas full-time and part-time employed patients were slightly less likely to participate. Time post-stroke at admission and length of stay were significantly associated with participation in cardiorespiratory exercise, with participants having a slightly longer time since stroke at admission (54.4 vs. 53.1 days, p = 0.010) and a longer rehabilitation stay (60.4 vs. 47.0 days, p < 0.001) compared with non-participants.

**Table 3.** Participant characteristics. Values presented are means with standard deviations in parentheses for continuous variables, or counts with percentages in parentheses for categorical variables.

| Participant characteristics | All patients | Participated in cardiorespiratory exercise | Did not participate in cardiorespiratory exercise | p-values |
| --- | --- | --- | --- | --- |
| Age (years) | 67.9 (14.7) | 68.2 (14.2) | 67.6 (15.0) | 0.47 |
| Sex (number) |  |  |  | 0.58 |
| Male | 870 (58) | 355 (59) | 515 (57) |  |
| Female | 627 (42) | 246 (41) | 381 (43) |  |
| Marital status (number) |  |  |  | 0.54 |
| Living single | 205 (14) | 85 (14) | 120 (13) |  |
| Living with a partner | 926 (62) | 369 (61) | 557 (62) |  |
| Widowed | 208 (14) | 77 (13) | 131 (15) |  |
| Separated | 158 (11) | 70 (12) | 88 (10) |  |
| Employment status (number) |  |  |  | 0.025 |
| Employed full-time | 475 (32) | 183 (30) | 292 (33) |  |
| Employed part-time | 83 (6) | 27 (4) | 56 (6) |  |
| Retired | 811 (54) | 351 (58) | 460 (51) |  |
| Unemployed | 81 (5) | 23 (4) | 58 (6) |  |
| Long-term disability | 44 (3) | 15 (2) | 29 (3) |  |
| Other | 3 (0) | 2 (0) | 1 (0) |  |
| Time post- stroke at admission (days) | 53.6 (78.2) | 54.4 (66.1) | 53.1 (85.4) | 0.010 |
| Length of stay (days) | 52.4 (44.2) | 60.4 (48.6) | 47.0 (40.2) | <0.001 |

### Main results

Among 1,497 patients who were undergoing rehabilitation after stroke, 601 (40.1%) participated in cardiorespiratory exercise (Table 2); 395 (26.4%) had cardiorespiratory exercise formally prescribed as part of their treatment plan, while 206 (13.8%) participated in incidental cardiorespiratory exercise that was not part of their treatment plan (Table 2). The total number of patients who participated in cardiorespiratory exercise during stroke rehabilitation was 94/271 (34.7%) before and 507/1226 (41.4%) after the onset of the COVID-19 pandemic (Table 2). Among these, 55/271 patients (20.3%) and 340/1226 patients (27.8%) had cardiorespiratory exercise included in their treatment plans before and after the onset of the pandemic respectively, whereas 39/271 patients (14.4%) and 167/1226 patients (13.6%) completed cardiorespiratory exercise that was not part of their treatment plans (Table 2).

Presence of a diagnosed cardiac disease (odds ratio (OR) = 0.74), poor left ventricular function (OR = 0.09), presence of a mental health condition (OR = 0.69), lower functional ambulation ability (OR = 0.74), and use of a wheelchair at admission to stroke rehabilitation (both indoors and outdoors (OR = 0.46), only when outdoors (OR = 0.35)), were significantly associated with lower participation in cardiorespiratory exercise after stroke (p-values < 0.05; Table 4). Use of a walker or rollator at admission to stroke rehabilitation (both indoors and outdoors (OR = 3.22), only when outdoors (OR = 3.89)), having a cardiorespiratory exercise relevant goal (OR = 2.13), and a longer length of stay for rehabilitation (OR = 1.01), were significantly associated with higher participation in cardiorespiratory exercise after stroke (p-values < 0.05; Table 4).

**Table 4.** Predictors of cardiorespiratory exercise participation during in-patient and out-patient stroke rehabilitation. Values are presented as counts with percentages in parentheses for categorical variables or means with percentages in parenthesis for continuous variables, and odds ratios (OR) with their corresponding 95% confidence intervals (CI) in brackets from multivariate logistic regression. OR: Odds Ratio; CI: Confidence Interval; N: Number; VIF: Variance Inflation Factor; FAC: Functional Ambulation Category; FIM: Functional Independence Measure;

| Participant characteristics | Participated in cardiorespiratory exercise (n=601) | Did not participate in cardiorespiratory exercise (n=896) | OR [95%CI] | p-value | VIF |
| --- | --- | --- | --- | --- | --- |
| Cardiac disease | 115 (19) | 214 (24) | 0.74 [0.56, 0.98] | 0.034 | 1.01 |
| Poor left ventricular function | 1 (<1) | 16 (2) | 0.09 [<1, 0.50] | 0.026 | 1.00 |
| Mental health condition | 91 (15) | 187 (21) | 0.69 [0.51, 0.93] | 0.015 | 1.00 |
| FAC (score) | 3.1 (1.6) | 3.3 (1.6) | 0.74 [0.66, 0.83] | <0.001 | 1.60 |
| Use of a walker or rollator at admission |  |  |  |  |  |
| No aid required | 229 (38) | 497 (55) | Reference |  | 1.12 |
| Only when indoors | 43 (7) | 152 (17) | 0.70 [0.44, 1.09] | 0.12 |  |
| Only when outdoors | 59 (10) | 33 (4) | 3.89 [2.44, 6.30] | <0.001 |  |
| Indoors and outdoors | 228 (38) | 146 (16) | 3.22 [2.40, 4.33] | <0.001 |  |
| Non-ambulatory | 42 (7) | 68 (8) | 0.86 [0.47, 1.54] | 0.61 |  |
| Use of a wheelchair at admission |  |  |  |  |  |
| No | 483 (80) | 627 (70) | Reference |  | 1.14 |
| Only when indoors | 12 (2) | 25 (3) | 0.79 [0.34, 1.73] | 0.56 |  |
| Only when outdoors | 13 (2) | 45 (5) | 0.35 [0.17, 0.68] | 0.003 |  |
| Indoors and outdoors | 93 (15) | 199 (22) | 0.46 [0.31, 0.69] | <0.001 |  |
| Having a cardiorespiratory fitness goal | 168 (28) | 146 (16) | 2.13 [1.61, 2.82] | <0.001 | 1.04 |
| Length of stay (days) | 60.4 (48.6) | 47.0 (40.2) | 1.01 [1.01, 1.01] | <0.001 | 1.04 |

The prediction performance of the full sample model resulted in a mean AUROC of 0.732 (standard deviation (SD) = 0.041), indicating moderate ability to distinguish between participants and non-participants of cardiorespiratory exercise during stroke rehabilitation. Sensitivity and specificity were both 0.675 (SD = 0.037), demonstrating balanced classification ability of the model.

In the inpatient subgroup model, predictors were younger age at admission (OR = 0.98); absence of a diagnosed cardiac disease (OR = 0.65); absence of mental health conditions (OR = 0.54); use of a walker or rollator at admission to stroke rehabilitation (both indoors and outdoors (OR = 6.16), only when outdoors (OR = 5.23)); not using wheelchair at admission to stroke rehabilitation (both indoors and outdoors (OR = 0.54), only when outdoors (OR = 0.15)); longer lengths of stay (OR = 1.02); and lower total FIM scores at admission (OR = 0.98) (Table 5).

**Table 5.** Predictors of cardiorespiratory exercise participation during inpatient stroke rehabilitation only. Values are presented as counts with percentages in parentheses for categorical variables or means with percentages in parenthesis for continuous variables, and odds ratios (OR) with their corresponding 95% confidence intervals (CI) in brackets from multivariate logistic regression. OR: Odds Ratio; CI: Confidence Interval; N: Number; VIF: Variance Inflation Factor; FAC: Functional Ambulation Category; FIM: Functional Independence Measure;

| Participant characteristics | Participated in cardiorespiratory exercise (n=299) | Did not participate in cardiorespiratory exercise (n=473) | OR[95%CI] | p-value | VIF |
| --- | --- | --- | --- | --- | --- |
| Age (years) | 70.3 (13.9) | 71.3 (14.2) | 0.98 [0.97, 1.00] | 0.009 | 1.08 |
| Cardiac disease | 61 (20) | 131 (28) | 0.66 [0.44, 0.98] | 0.041 | 1.02 |
| Mental health condition | 47 (16) | 108 (23) | 0.57 [0.37, 0.88] | 0.012 | 1.01 |
| Use of a walker or rollator at admission |  |  |  |  | 1.10 |
| No aid required | 56 (19) | 187 (40) | Reference |  |  |
| Only when indoors | 27 (9) | 130 (27) | 0.67 [0.36, 1.21] | 0.19 |  |
| Only when outdoors | 20 (7) | 15 (3) | 4.33 [1.99, 9.61] | <0.001 |  |
| Indoors and outdoors | 159 (53) | 83 (18) | 6.12 [4.01, 9.46] | <0.001 |  |
| Non-ambulatory | 37 (12) | 58 (12) | 1.10 [0.56, 2.13] | 0.78 |  |
| Use of a wheelchair at admission |  |  |  |  | 1.11 |
| No | 218 (73) | 258 (55) | Reference |  |  |
| Only when indoors | 11 (4) | 20 (4) | 1.32 [0.51, 3.30] | 0.56 |  |
| Only when outdoors | 3 (1) | 30 (6) | 0.14 [0.03, 0.47] | 0.004 |  |
| Indoors and outdoors | 67 (22) | 165 (35) | 0.52 [0.32, 0.84] | 0.008 |  |
| Length of stay (days) | 34.6 (20.8) | 28.6 (18.0) | 1.02 [1.01, 1.03] | <0.001 | 1.31 |
| FIM total (score) | 65.8 (16.2) | 71.7 (19.2) | 0.98 [0.97, 0.99] | 0.003 | 1.38 |

The prediction performance of the inpatient subgroup model resulted in a mean AUROC of 0.778 (SD = 0.053), indicating moderate to good discriminative ability to distinguish between participants and non-participants of cardiorespiratory exercise during stroke rehabilitation. Sensitivity and specificity were both 0.701 (SD = 0.053), demonstrating balanced classification ability of the model.

The sensitivity analysis revealed a moderate to substantial improvement (5.5%) in AUROC (Δ = 0.054) in the model that included inpatients only (AUROC = 0.792) when compared to the full sample with both inpatients and outpatients (AUROC = 0.738), along with a difference in the predictors of participation in cardiorespiratory exercise during stroke rehabilitation between the full sample and the inpatient subgroup. The Hosmer-Lemeshow tests for models with all patients and inpatients only indicated no significant difference between the observed and expected frequencies, indicating a good fit (all patients: χ^2^ (8, N = 1497) = 9.56, p = 0.30; inpatients only: χ^2^ (8, N = 772) = 12.07, p = 0.15).

The majority (59%) of those who participated in cardiorespiratory exercise during stroke rehabilitation completed a 2-minute walk test; only about 2% underwent a standardized cardiopulmonary exercise test.^16,17^ The majority (81.5%) used a recumbent stepper, while others used a cycle ergometer (5%), a treadmill (5.2%), or no equipment (2.2%). Overall, only 1.5% of patients (N = 9/601) who participated in cardiorespiratory exercise during stroke rehabilitation did so at a dose that adhered to evidence-based guidelines, which recommend moderate-intensity exercise for at least 20 minutes per session (Table 6).^16,17^

**Table 6.**
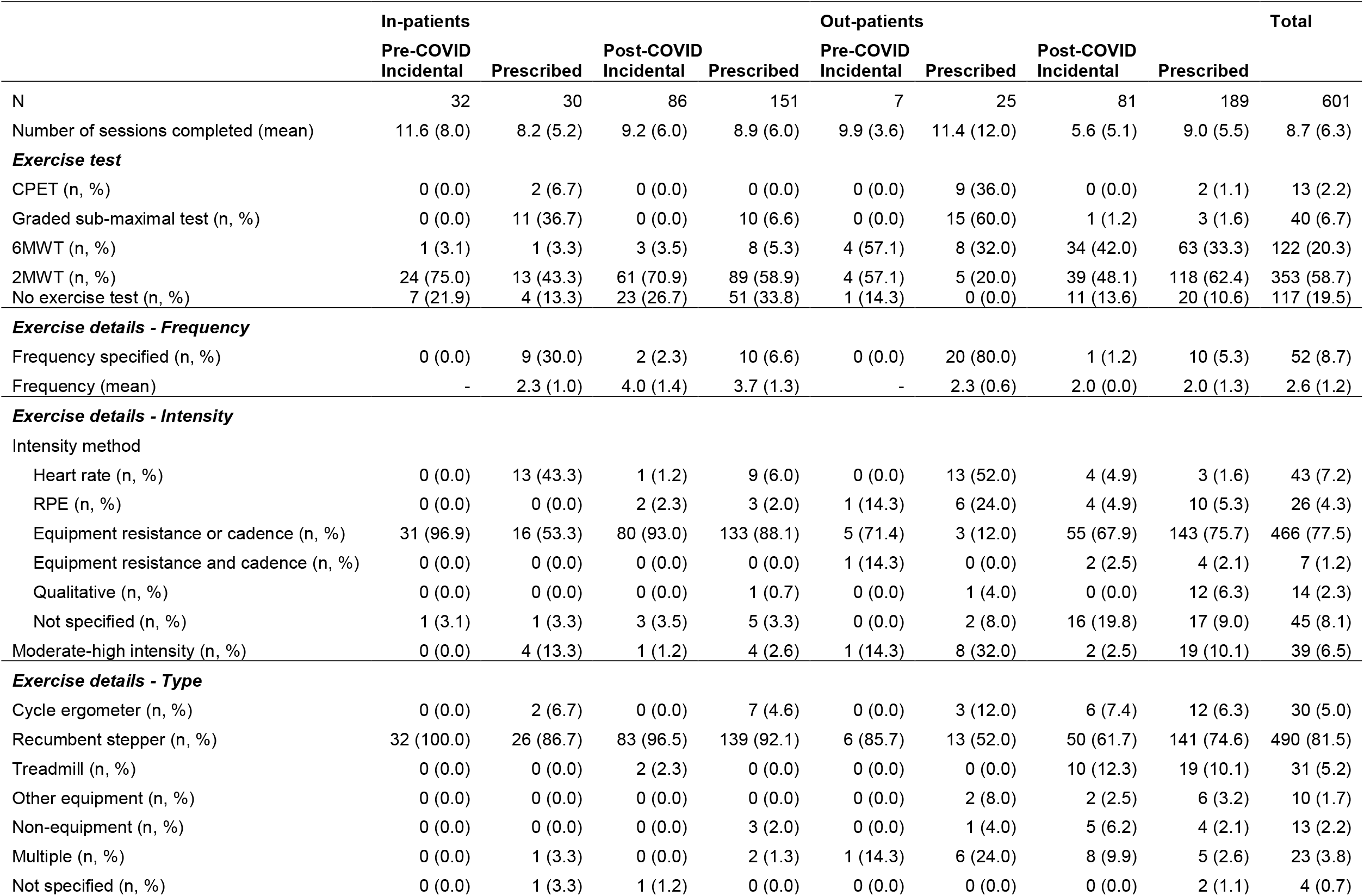
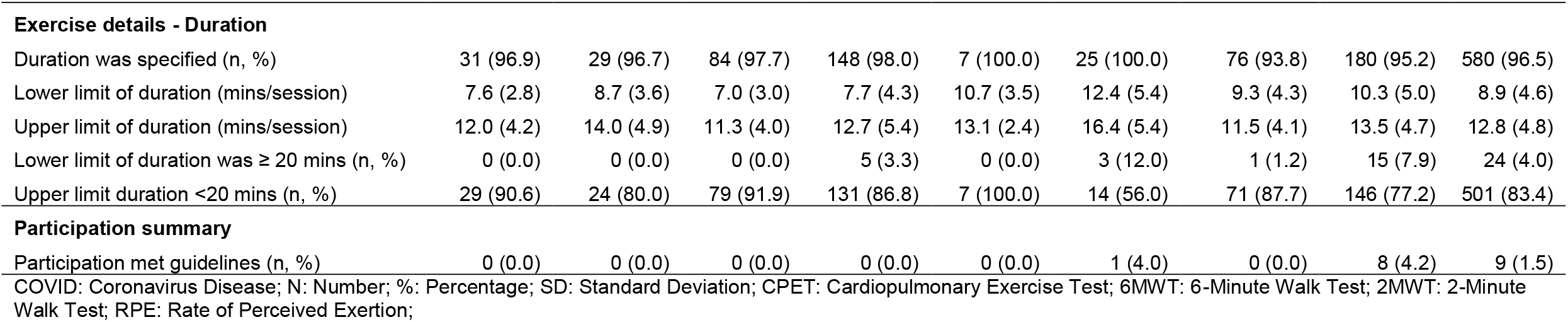
Exercise prescription details. Values presented are means with standard deviations in parentheses for continuous variables, or counts with percentages in parentheses for categorical variables. Exercise participation met guidelines if it was at least of moderate intensity and the lower limit of exercise duration was ≥ 20 minutes.

|  | In-patients |  |  |  | Out-patients |  |  |  | Total |
| --- | --- | --- | --- | --- | --- | --- | --- | --- | --- |
|  | Pre-COVID<br>Incidental | Prescribed | Post-COVID<br>Incidental | Prescribed | Pre-COVID<br>Incidental | Prescribed | Post-COVID<br>Incidental | Prescribed |  |
| N | 32 | 30 | 86 | 151 | 7 | 25 | 81 | 189 | 601 |
| Number of sessions completed (mean) | 11.6 (8.0) | 8.2 (5.2) | 9.2 (6.0) | 8.9 (6.0) | 9.9 (3.6) | 11.4 (12.0) | 5.6 (5.1) | 9.0 (5.5) | 8.7 (6.3) |
| **Exercise test** |  |  |  |  |  |  |  |  |  |
| CPET (n, %) | 0 (0.0) | 2 (6.7) | 0 (0.0) | 0 (0.0) | 0 (0.0) | 9 (36.0) | 0 (0.0) | 2 (1.1) | 13 (2.2) |
| Graded sub-maximal test (n, %) | 0 (0.0) | 11 (36.7) | 0 (0.0) | 10 (6.6) | 0 (0.0) | 15 (60.0) | 1 (1.2) | 3 (1.6) | 40 (6.7) |
| 6MWT (n, %) | 1 (3.1) | 1 (3.3) | 3 (3.5) | 8 (5.3) | 4 (57.1) | 8 (32.0) | 34 (42.0) | 63 (33.3) | 122 (20.3) |
| 2MWT (n, %) | 24 (75.0) | 13 (43.3) | 61 (70.9) | 89 (58.9) | 4 (57.1) | 5 (20.0) | 39 (48.1) | 118 (62.4) | 353 (58.7) |
| No exercise test (n, %) | 7 (21.9) | 4 (13.3) | 23 (26.7) | 51 (33.8) | 1 (14.3) | 0 (0.0) | 11 (13.6) | 20 (10.6) | 117 (19.5) |
| **Exercise details - Frequency** |  |  |  |  |  |  |  |  |  |
| Frequency specified (n, %) | 0 (0.0) | 9 (30.0) | 2 (2.3) | 10 (6.6) | 0 (0.0) | 20 (80.0) | 1 (1.2) | 10 (5.3) | 52 (8.7) |
| Frequency (mean) | - | 2.3 (1.0) | 4.0 (1.4) | 3.7 (1.3) | - | 2.3 (0.6) | 2.0 (0.0) | 2.0 (1.3) | 2.6 (1.2) |
| **Exercise details - Intensity** |  |  |  |  |  |  |  |  |  |
| Intensity method |  |  |  |  |  |  |  |  |  |
| Heart rate (n, %) | 0 (0.0) | 13 (43.3) | 1 (1.2) | 9 (6.0) | 0 (0.0) | 13 (52.0) | 4 (4.9) | 3 (1.6) | 43 (7.2) |
| RPE (n, %) | 0 (0.0) | 0 (0.0) | 2 (2.3) | 3 (2.0) | 1 (14.3) | 6 (24.0) | 4 (4.9) | 10 (5.3) | 26 (4.3) |
| Equipment resistance or cadence (n, %) | 31 (96.9) | 16 (53.3) | 80 (93.0) | 133 (88.1) | 5 (71.4) | 3 (12.0) | 55 (67.9) | 143 (75.7) | 466 (77.5) |
| Equipment resistance and cadence (n, %) | 0 (0.0) | 0 (0.0) | 0 (0.0) | 0 (0.0) | 1 (14.3) | 0 (0.0) | 2 (2.5) | 4 (2.1) | 7 (1.2) |
| Qualitative (n, %) | 0 (0.0) | 0 (0.0) | 0 (0.0) | 1 (0.7) | 0 (0.0) | 1 (4.0) | 0 (0.0) | 12 (6.3) | 14 (2.3) |
| Not specified (n, %) | 1 (3.1) | 1 (3.3) | 3 (3.5) | 5 (3.3) | 0 (0.0) | 2 (8.0) | 16 (19.8) | 17 (9.0) | 45 (8.1) |
| Moderate-high intensity (n, %) | 0 (0.0) | 4 (13.3) | 1 (1.2) | 4 (2.6) | 1 (14.3) | 8 (32.0) | 2 (2.5) | 19 (10.1) | 39 (6.5) |
| **Exercise details - Type** |  |  |  |  |  |  |  |  |  |
| Cycle ergometer (n, %) | 0 (0.0) | 2 (6.7) | 0 (0.0) | 7 (4.6) | 0 (0.0) | 3 (12.0) | 6 (7.4) | 12 (6.3) | 30 (5.0) |
| Recumbent stepper (n, %) | 32 (100.0) | 26 (86.7) | 83 (96.5) | 139 (92.1) | 6 (85.7) | 13 (52.0) | 50 (61.7) | 141 (74.6) | 490 (81.5) |
| Treadmill (n, %) | 0 (0.0) | 0 (0.0) | 2 (2.3) | 0 (0.0) | 0 (0.0) | 0 (0.0) | 10 (12.3) | 19 (10.1) | 31 (5.2) |
| Other equipment (n, %) | 0 (0.0) | 0 (0.0) | 0 (0.0) | 0 (0.0) | 0 (0.0) | 2 (8.0) | 2 (2.5) | 6 (3.2) | 10 (1.7) |
| Non-equipment (n, %) | 0 (0.0) | 0 (0.0) | 0 (0.0) | 3 (2.0) | 0 (0.0) | 1 (4.0) | 5 (6.2) | 4 (2.1) | 13 (2.2) |
| Multiple (n, %) | 0 (0.0) | 1 (3.3) | 0 (0.0) | 2 (1.3) | 1 (14.3) | 6 (24.0) | 8 (9.9) | 5 (2.6) | 23 (3.8) |
| Not specified (n, %) | 0 (0.0) | 1 (3.3) | 1 (1.2) | 0 (0.0) | 0 (0.0) | 0 (0.0) | 0 (0.0) | 2 (1.1) | 4 (0.7) |
|  | Pre-COVID<br>Incidental | Prescribed | Post-COVID<br>Incidental | Prescribed | Pre-COVID<br>Incidental | Prescribed | Post-COVID<br>Incidental | Prescribed |  |
| Exercise details - Duration |  |  |  |  |  |  |  |  |  |
| Duration was specified (n, %) | 31 (96.9) | 29 (96.7) | 84 (97.7) | 148 (98.0) | 7 (100.0) | 25 (100.0) | 76 (93.8) | 180 (95.2) | 580 (96.5) |
| Lower limit of duration (mins/session) | 7.6 (2.8) | 8.7 (3.6) | 7.0 (3.0) | 7.7 (4.3) | 10.7 (3.5) | 12.4 (5.4) | 9.3 (4.3) | 10.3 (5.0) | 8.9 (4.6) |
| Upper limit of duration (mins/session) | 12.0 (4.2) | 14.0 (4.9) | 11.3 (4.0) | 12.7 (5.4) | 13.1 (2.4) | 16.4 (5.4) | 11.5 (4.1) | 13.5 (4.7) | 12.8 (4.8) |
| Lower limit of duration was ≥ 20 mins (n, %) | 0 (0.0) | 0 (0.0) | 0 (0.0) | 5 (3.3) | 0 (0.0) | 3 (12.0) | 1 (1.2) | 15 (7.9) | 24 (4.0) |
| Upper limit duration <20 mins (n, %) | 29 (90.6) | 24 (80.0) | 79 (91.9) | 131 (86.8) | 7 (100.0) | 14 (56.0) | 71 (87.7) | 146 (77.2) | 501 (83.4) |
| Participation summary |  |  |  |  |  |  |  |  |  |
| Participation met guidelines (n, %) | 0 (0.0) | 0 (0.0) | 0 (0.0) | 0 (0.0) | 0 (0.0) | 1 (4.0) | 0 (0.0) | 8 (4.2) | 9 (1.5) |

COVID: Coronavirus Disease; N: Number; %: Percentage; SD: Standard Deviation; CPET: Cardiopulmonary Exercise Test; 6MWT: 6-Minute Walk Test; 2MWT: 2-Minute Walk Test; RPE: Rate of Perceived Exertion;

## DISCUSSION

Approximately 40% of stroke patients undergoing rehabilitation participated in cardiorespiratory exercise. Lower rates of participation were associated with diagnosed cardiac disease, poor left ventricular function, presence of mental health conditions, reduced ambulation ability, and wheelchair use at rehabilitation admission. Higher rates of participation were associated with using a walker or rollator at admission, having a specific goal to engage in cardiorespiratory exercise, and a longer rehabilitation stay. Only 1.5% of those who completed cardiorespiratory exercise met evidence-based guidelines, which recommend at least 20 minutes of moderate-intensity exercise per session).^16,17^

While 40% participated in cardiorespiratory exercise, only 26% took part in a structured program through a formal prescription. The relatively high proportion of patients participating incidentally suggests that there may be potential barriers to formal prescription, such as limited resources to conduct safe exercise testing, mobility barriers, or patient preference.^26,27^ This highlights missed opportunities for structured exercise programming within rehabilitation plans, which require further investigation and targeted strategies. Participation rates were overall slightly lower prior to the COVID-19 pandemic; therefore, the low rates of exercise participation in the current study cannot be explained by changes in care delivery as a result of the pandemic.

Diagnosed cardiac disease and left ventricular dysfunction were associated with lower participation in cardiorespiratory exercise during stroke rehabilitation. Left ventricular dysfunction is a hallmark feature of heart failure which can present as either asymptomatic or symptomatic disease, with some individuals exhibiting structural cardiac abnormalities without overt clinical symptoms.^28^ Symptom-limited or submaximal cardiopulmonary exercise testing with electrocardiography is recommended for individuals with asymptomatic heart failure to ensure safe aerobic training prescription post-stroke;^29^ however, this equipment or testing resource were often unavailable to physiotherapists. Since exercise is contraindicated for symptomatic patients, such as those with New York Heart Association Class IV heart failure,^30^ it was likely appropriate to exclude individuals with left ventricular dysfunction from moderate-to-vigorous exercise programs.^29^ In our chart review, we lacked information regarding the severity or acuity of medical conditions, which may be a factor in the decision not to prescribe cardiorespiratory exercise post-stroke. However, patients with left ventricular dysfunction due to a cardiac disease such as heart failure have shown improvements in cardiac remodeling following cardiorespiratory exercise, as demonstrated by reductions in left ventricular end-diastolic diameter.^31,32^ Conversely, patients with left ventricular dysfunction resulting from non-cardiac critical illness, such as sepsis, respiratory failure, major loss of blood, and neurological disorders such as status epilepticus, traumatic brain injury, and subarachnoid haemorrhage, often present with regional hypokinesia, which in turn is linked to a higher risk of mortality.^33^ It remains unclear whether patients who develop left ventricular dysfunction due to non-cardiac critical illness are at risk of adverse events from cardiorespiratory exercise during stroke rehabilitation. Therefore, further evaluation of patients with these conditions is necessary, as they may represent a distinct cohort requiring tailored rehabilitation strategies. For example, if a gradual progression of cardiorespiratory exercise during stroke rehabilitation can improve ejection fraction over time more than its natural history with medical therapy alone, this may define a unique subgroup of stroke patients who respond favorably to exercise-based interventions.^34^

About 23% of patients who did not participate in cardiorespiratory exercise after stroke had a diagnosed mental health condition, compared to 16% of those who participated, suggesting that the presence of mental health conditions may be a barrier to participation in exercise-based rehabilitation after stroke. Evidence suggests that screening for mental health conditions at the time of admission into stroke rehabilitation,^35^ incorporating mental health support, such as counselling, to improve exercise participation,^36^ and referral to multidisciplinary interventions to provide personalized support,^37^ may improve self-directed participation in cardiorespiratory exercise after stroke. Addressing mental health concerns may therefore be important for optimizing participation in cardiorespiratory exercise following stroke.

Patients with reduced ambulation ability or those requiring a wheelchair at admission were less likely to engage in cardiorespiratory exercise, possibly due to mobility limitations or perceived safety concerns. Conversely, individuals who used a walker or rollator at admission had higher participation rates, suggesting that a certain level of independent mobility may facilitate engagement in cardiorespiratory exercise. Wong et al.^38^ reported that wheelchair users increased their participation in cardiorespiratory exercise when provided with adapted equipment, which was perceived as both usable and enjoyable. In this study, most participants used a recumbent cross-trainer, which is suitable for individuals with severe mobility impairments; however, participation may still have been limited by practical barriers, such as the need for a heavy transfer from the wheelchair to the recumbent cross-trainer, indicating that factors beyond physical capacity may have influenced engagement in cardiorespiratory exercise programs. Among stroke rehabilitation programs that included cardiorespiratory exercise component, most (n = 27/36; 75%) reported excluding those with severe mobility deficits.^9^ This is concerning because individuals with severe mobility deficits require interventions to overcome the barriers to participation, not exclusion from cardiorespiratory exercise due to their disability.^39^ Similar findings have been reported for other patient populations; in a recent survey, only 48% of cardiac rehabilitation programs accepted referrals for people with lower limb amputations and severe mobility deficits.^40^ Similarly, only 40.6% accepted referrals for those with peripheral arterial disease and severe mobility limitations, such as critical limb ischemia.^41^ It is possible that individuals with moderate mobility limitations were more likely to receive extended rehabilitation than those at either end of the functional mobility range, highly ambulatory individuals who may recover more quickly and be discharged earlier, and non-ambulatory individuals who may be transferred to institutional care when the potential to benefit from intensive rehabilitation is limited.^42^ These findings highlight the importance of improving rehabilitation planning across the continuum of care, ensuring that opportunities to participate in aerobic exercise extend beyond discrete inpatient admission criteria and are supported through coordinated transitions from inpatient to outpatient and community-based rehabilitation services.^42^

Only 1.5% of stroke rehabilitation patients in our study met evidence-based guidelines for cardiorespiratory exercise,^16,17^ which recommend at least 20 minutes of moderate-intensity exercise per session. Therefore, our study found that physiotherapists did not consistently prescribe intensity and duration in accordance with stroke rehabilitation guidelines, highlighting a significant gap between recommended practice and implementation in clinical settings. Previous studies have highlighted similar challenges in clinical practice, particularly in delivering the recommended exercise dosage to patients with stroke. For instance, Billinger et al.^16^ noted that although moderate-intensity aerobic exercise is important for improving cardiorespiratory fitness and promoting neuroplasticity post-stroke, various patient-related factors, such as fatigue, lack of motivation, and fear (of falling, subsequent stroke, and other adverse events), often affect adherence. Additionally, Girard et al.^43^ reported that even within structured stroke rehabilitation programs, consistently achieving the recommended exercise dosage necessary to elicit a cardiopulmonary training effect and support the physiological demands of independent living remains challenging. We found that stroke rehabilitation patients exercised for an average of 12.8 minutes per session, and over 80% had their sessions limited to less than 20 minutes. In our study, progression in exercise prescription may have been limited by factors such as clinical resource constraints (including limited duration of therapy sessions), safety concerns for medically complex patients, and the lack of standardized exercise tests to guide and monitor progression, highlighting the need for structured, evidence-based approaches to support dose progression in cardiorespiratory training during stroke rehabilitation.^44^

### Limitations

While certain factors were associated with reduced participation in cardiorespiratory exercise, causality cannot be established. The absence of data on the acuity or severity of cardiac conditions further limits interpretation, as some patients might have had exercise restrictions due to clinical contraindications. Other limitations include the potential for missing or incomplete information, non-standardized recording practices, and unreported information in the medical records. The COVID-19 pandemic may have further influenced rehabilitation practices,^45^ potentially affecting both the recruitment of patients and adherence to cardiorespiratory exercise programs. These issues could have limited the representativeness of our findings, impacting the generalizability of our results. However, we included all patients admitted for stroke rehabilitation during the study period to avoid selection bias. Lastly, the results from the four Canadian stroke rehabilitation centres may not apply to other healthcare settings in Canada or internationally.

## Data Availability

Raw data are not available publicly due to local privacy legislation.

## Acknowledgments

We thank Kay-Ann Allen, Cynthia Danells, Hanna Fang, David Jagroop and Alex Kalli for their assistance with data collection.

## Ethical approval and informed consent statements

Research ethics approval was obtained from the research ethics boards of the University Health Network (protocol number: 20-5695), Sunnybrook Health Sciences Centre (protocol number: 3605), and Hamilton Integrated Research Ethics Board (protocol number: 11523), and a waiver of informed consent for chart review was granted.

---

**Supplementary Material A: Summary of missing variables**

| **Variable Names** | **Missing (n)** |
| --- | --- |
| Marital Status | 88 |
| Employment Status | 65 |
| FIM Total | 844 |
| FIM Motor | 850 |
| FIM Cognitive | 851 |
| FAC Score | 245 |
| Use of Cane | 63 |
| Use of Walker | 64 |
| Use of Wheelchair | 81 |
| MoCA Total | 783 |
| NIHSS Scale Score | 1193 |
| Having A Cardiorespiratory Exercise Goal | 31 |
| Time Post Stroke (days) | 10 |
| Length Of Stay in Rehabilitation (days) | 3 |
| CMSA Arm | 731 |
| CMSA Hand | 734 |
| CMSA Leg | 808 |
| CMSA Foot | 816 |

FIM: functional independence measure; FAC: functional ambulation category; MoCA: Montreal Cognitive Assessment; NIHSS: National Institutes of Health Stroke Severity; CMSA: Chedoke McMaster Stroke Assessment;

## REFERENCES

1. MacKay-Lyons MJ, Makrides L. Exercise capacity early after stroke. Archives of Physical Medicine and Rehabilitation 2002; 83: 1697–1702.

2. Tang A, Sibley KM, Thomas SG, et al. Maximal Exercise Test Results in Subacute Stroke. Archives of Physical Medicine and Rehabilitation 2006; 87: 1100–1105.

3. Rimmer JH, Wang E. Aerobic Exercise Training in Stroke Survivors. Topics in Stroke Rehabilitation 2005; 12: 17–30.

4. Caspersen CJ, Powell KE, Christenson GM. Physical activity, exercise, and physical fitness: definitions and distinctions for health-related research. Public Health Rep 1985; 100: 126–131.

5. Ross R, Blair SN, Arena R, et al. Importance of Assessing Cardiorespiratory Fitness in Clinical Practice: A Case for Fitness as a Clinical Vital Sign: A Scientific Statement From the American Heart Association. Circulation; 134. Epub ahead of print 13 December 2016. DOI: 10.1161/CIR.0000000000000461.

6. Billinger SA, Coughenour E, MacKay-Lyons MJ, et al. Reduced Cardiorespiratory Fitness after Stroke: Biological Consequences and Exercise-Induced Adaptations. Stroke Research and Treatment 2012; 2012: 1–11.

7. Prout EC, Brooks D, Mansfield A, et al. Patient Characteristics That Influence Enrollment and Attendance in Aerobic Exercise Early After Stroke. Archives of Physical Medicine and Rehabilitation 2015; 96: 823–830.

8. Biasin L, Sage MD, Brunton K, et al. Integrating Aerobic Training Within Subacute Stroke Rehabilitation: A Feasibility Study. Physical Therapy 2014; 94: 1796–1806.

9. Nathoo C, Buren S, El-Haddad R, et al. Aerobic Training in Canadian Stroke Rehabilitation Programs. Journal of Neurologic Physical Therapy 2018; 42: 248–255.

10. Doyle L, MacKay-Lyons M. Utilization of Aerobic Exercise in Adult Neurological Rehabilitation by Physical Therapists in Canada. Journal of Neurologic Physical Therapy 2013; 37: 20–26.

11. Mansfield A, Brooks D, Tang A, et al. Promoting Optimal Physical Exercise for Life (PROPEL): aerobic exercise and self-management early after stroke to increase daily physical activity—study protocol for a stepped-wedge randomised trial. BMJ Open 2017; 7: e015843.

12. Gaskins NJ, Bray E, Hill JE, et al. Factors influencing implementation of aerobic exercise after stroke: a systematic review. Disability and Rehabilitation 2021; 43: 2382–2396.

13. Moncion K, Biasin L, Jagroop D, et al. Barriers and Facilitators to Aerobic Exercise Implementation in Stroke Rehabilitation: A Scoping Review. Journal of Neurologic Physical Therapy 2020; 44: 179–187.

14. Barzideh A, Devasahayam AJ, Tang A, et al. Physiotherapists’ use of aerobic exercise during stroke rehabilitation: a qualitative study using chart-stimulated recall. Disability and Rehabilitation 2025; 1–13.

15. Prout EC, Mansfield A, McIlroy WE, et al. Physiotherapists’ perspectives on aerobic exercise early after stroke: A preliminary study. Physiotherapy Theory and Practice 2016; 32: 452–460.

16. Billinger SA, Arena R, Bernhardt J, et al. Physical Activity and Exercise Recommendations for Stroke Survivors: A Statement for Healthcare Professionals From the American Heart Association/American Stroke Association. Stroke 2014; 45: 2532–2553.

17. MacKay-Lyons M, Billinger SA, Eng JJ, et al. Aerobic Exercise Recommendations to Optimize Best Practices in Care After Stroke: AEROBICS 2019 Update. Physical Therapy 2020; 100: 149– 156.

18. Vandenbroucke JP, Von Elm E, Altman DG, et al. Strengthening the Reporting of Observational Studies in Epidemiology (STROBE): Explanation and Elaboration. PLoS Med 2007; 4: e297.

19. Keith RA, Granger CV, Hamilton BB, et al. The functional independence measure: a new tool for rehabilitation. Adv Clin Rehabil 1987; 1: 6–18.

20. Holden MK, Gill KM, Magliozzi MR, et al. Clinical Gait Assessment in the Neurologically Impaired. Physical Therapy 1984; 64: 35–40.

21. Gowland C, Stratford P, Ward M, et al. Measuring physical impairment and disability with the Chedoke-McMaster Stroke Assessment. Stroke 1993; 24: 58–63.

22. Nasreddine ZS, Phillips NA, Bédirian V, et al. The Montreal Cognitive Assessment, MoCA: A Brief Screening Tool For Mild Cognitive Impairment. J American Geriatrics Society 2005; 53: 695–699.

23. Goldstein LB, Bertels C, Davis JN. Interrater Reliability of the NIH Stroke Scale. Archives of Neurology 1989; 46: 660–662.

24. Borg G. Borg’s Perceived exertion and pain scales. Champaign, IL: Human Kinetics, 1998.

25. Pescatello LS, American College of Sports Medicine (eds). ACSM’s guidelines for exercise testing and prescription. 9. ed. Philadelphia: Wolters Kluwer/Lippincott Williams & Wilkins, 2014.

26. Inness EL, Aqui A, Foster E, et al. Determining Safe Participation in Aerobic Exercise Early After Stroke Through a Graded Submaximal Exercise Test. Physical Therapy 2020; 100: 1434– 1443.

27. Regan EW, Handlery R, Stewart JC, et al. Feasibility of integrating survivors of stroke into cardiac rehabilitation: A mixed methods pilot study. PLoS ONE 2021; 16: e0247178.

28. Heidenreich PA, Bozkurt B, Aguilar D, et al. 2022 AHA/ACC/HFSA Guideline for the Management of Heart Failure. Journal of the American College of Cardiology 2022; 79: e263– e421.

29. Inness E, Jagroop D, Biasin L, et al. The STroke AeRobic exercise implementation Toolkit-START. Sage Publications Ltd 1 Olivers Yard, 55 City Road, London EC1Y 1SP, England, 2024, pp. 23–24.

30. Taylor JL, Myers J, Bonikowske AR. Practical guidelines for exercise prescription in patients with chronic heart failure. Heart Fail Rev 2023; 28: 1285–1296.

31. Qi Z, Zheng Y, Chan JSK, et al. Exercise-based cardiac rehabilitation for left ventricular function in patients with heart failure: A systematic review and meta-analysis. Current Problems in Cardiology 2024; 49: 102210.

32. Tucker WJ, Beaudry RI, Liang Y, et al. Meta-analysis of Exercise Training on Left Ventricular Ejection Fraction in Heart Failure with Reduced Ejection Fraction: A 10-year Update. Prog Cardiovasc Dis 2019; 62: 163–171.

33. Cavefors O, Holmqvist J, Bech-Hanssen O, et al. Regional left ventricular systolic dysfunction associated with critical illness: incidence and effect on outcome. ESC Heart Fail 2021; 8: 5415– 5423.

34. Behrman B, Aronow WS, Frishman WH. Recovery From Left Ventricular Dysfunction. Cardiology in Review 2024; 32: 408–416.

35. Tjokrowijoto P, Kneebone I, Baker C, et al. Mental health support after stroke: A qualitative exploration of lived experience. Rehabilitation Psychology 2024; 69: 195–205.

36. Lanctôt KL, Lindsay MP, Smith EE, et al. Canadian Stroke Best Practice Recommendations: Mood, Cognition and Fatigue following Stroke, 6th edition update 2019. International Journal of Stroke 2020; 15: 668–688.

37. Wen X, Li Y, Zhang Q, et al. Enhancing long-term adherence in elderly stroke rehabilitation through a digital health approach based on multimodal feedback and personalized intervention. Sci Rep; 15. Epub ahead of print 23 April 2025. DOI: 10.1038/s41598-025-95726-z.

38. Wong RN, Stewart AL, Sawatzky B, et al. Exploring exercise participation and the usability of the adaptive rower and arm crank ergometer through wheelchair users’ perspectives. Disability and Rehabilitation 2022; 44: 3915–3924.

39. Toma J, Hammond B, Chan V, et al. Inclusion of People Poststroke in Cardiac Rehabilitation Programs in Canada: A Missed Opportunity for Referral. CJC Open 2020; 2: 195–206.

40. Marzolini S, Brunne A, Hébert A-A, et al. Barriers and Facilitators to Cardiovascular Rehabilitation Programmes for People with Lower Limb Amputation: A Survey of Clinical Practice in Canada. Physiother Can 2024; 76: 199–208.

41. Ahden S, Ngo V, Hoskin J, et al. Inclusion of People With Peripheral Artery Disease in Cardiac Rehabilitation Programs: A Pan-Canadian Survey. Heart Lung Circ 2021; 30: 1031–1043.

42. Janzen S, Mirkowski M, McIntyre A, et al. Referral patterns of stroke rehabilitation inpatients to a model system of outpatient services in Ontario, Canada: a 7-year retrospective analysis. BMC Health Serv Res 2019; 19: 399.

43. Girard V, Bellavance-Tremblay H, Gaudet-Drouin G, et al. Cardiorespiratory strain during stroke rehabilitation: Are patients trained enough? A systematic review. Ann Phys Rehabil Med 2021; 64: 101443.

44. Inness EL, Jagroop D, Andreoli A, et al. Factors That Influence the Clinical Implementation of Aerobic Exercise in Stroke Rehabilitation: A Theory-Informed Qualitative Study. Physical Therapy 2022; 102: pzac014.

45. Salbach NM, Mountain A, Lindsay MP, et al. Canadian Stroke Best Practice Recommendations: Virtual Stroke Rehabilitation Interim Consensus Statement 2022. Am J Phys Med Rehabil 2022; 101: 1076–1082.

